# Dengue virus in Solomon Islands 2023-2025: a whole genome surveillance study

**DOI:** 10.64898/2026.06.30.26356797

**Authors:** Jean Moselen, Eike Steinig, Andrew Darcy, Alfred Dofai, Angella Manele, Brenda Lauri, Cynthia Joshua, Paul Mauruwai, Ammar Aziz, Paul F. Horwood, Nicole Orlando, Leon Caly, Navin Karan, Janella Solomon, Chuan Kok Lim

## Abstract

**Background:** Following the cessation of COVID-19 travel restrictions in July 2022, concerns about a delayed dengue outbreak prompted the Solomon Islands Ministry of Health to establish enhanced genomic surveillance of circulating dengue virus (DENV) strains.

**Methods:** We performed amplicon-based whole genome sequencing (WGS) on PCR-positive serum samples collected at the National Referral Hospital, Honiara, between January 2023 and March 2025 (n = 63). Genomes were compared with publicly available sequences, and maximum-likelihood phylogenies were used to explore regional transmission dynamics.

**Findings:** We generated the first whole genome sequences from Solomon Islands (n = 45), with high recovery rates from acute infections (90%, Ct 17-44, mean coverage > 80%). Co-circulation of DENV-1, DENV-2, and DENV-4 was observed, with evidence of a serotype shift emerging in 2024. Phylogeographic analyses suggest ancestral introductions from Papua New Guinea for DENV-1 and DENV-2.

**Interpretation:** This study demonstrates the feasibility of whole genome sequencing for dengue surveillance in Solomon Islands through a referral sequencing model that provides a pathway for progressive local capacity-building. By addressing technical challenges and critical gaps in regional genomic representation, our findings strengthen the evidence base needed for equitable and sustainable implementation of pathogen genomics across Pacific Island countries and territories.

**Funding:** This work was supported by the Victorian Infectious Diseases Reference Laboratory (VIDRL).

## Introduction

Dengue virus (DENV) is the most prevalent arthropod-borne pathogen globally, causing an estimated 390 million infections and 96 million clinical cases each year [1]. It is classified into four serogroups (DENV-1 to DENV-4), which are endemic across 128 countries and over-represented in tropical and sub-tropical climates [2, 3]. Dengue virus is the leading cause of arboviral disease in the Western Pacific region, with outbreaks having been documented since the 19th century [4]. This region spans from central Asia to Oceania and is comprised of 38 countries and areas with a population of 2.2 billion people. The Western Pacific region includes 22 Pacific Island countries in which 19 have reported ≥ 10 Dengue cases in ≥ 3 of the past ten years [4, 5]. Co-circulation of all DENV serotypes (DENV-1-4) in the Western Pacific region have been reported over the last decade, each with varying magnitude, duration and severity [4]. However, transmission dynamics of DENV in many Pacific Island countries remain poorly understood, making it challenging to assess the burden of the disease and the potential impact of control interventions [6]. Tropical climate conditions in Pacific Island countries and territories contribute to high mosquito prevalence and result in hyper-endemic, year-round DENV transmission [7]. Local case surveillance and consistent reporting are essential for effective outbreak response and mitigation [8] but the implementation and perpetuation of arthropod-borne surveillance measures can be challenging for many small island nations, including Solomon Islands [9, 10].

Dengue activity in Solomon Islands is notable for its cyclical serotype displacement dynamics, with a single dominant DENV serotype circulating for three to five years before being rapidly displaced by another [6, 11]. Documented displacement events include the replacement of DENV-1 by DENV-4 in 2007 [12], followed by DENV-3 in 2013 [8, 13] and DENV-2 in 2016 [6]. Solomon Islands experienced an unprecedented DENV-2 outbreak in 2016-2017 [3, 9, 11, 13] where 12,329 suspected cases were reported across 9 of the country’s 10 provinces, including 877 hospitalisations, 1510 laboratory confirmed cases, and 16 deaths. Dengue surveillance initiatives were impacted during the COVID-19 pandemic from March 2020 until travel restrictions were lifted in July 2022. Following the reopening of international borders, the Solomon Islands Ministry of Health highlighted public health risks from reduced Dengue surveillance, including undetected serotype displacement and the potential emergence of an outbreak like that in 2016. In response, an enhanced surveillance program was implemented in Honiara to monitor circulating strains. While such measures are essential for timely outbreak detection and response, sustaining robust surveillance remains challenging in resource-limited Pacific Island settings. Strengthening these systems is critical not only for mitigating local dengue epidemics but also for understanding serotype dynamics that shape transmission risk across the wider Western Pacific region.

Molecular characterisation of dengue virus (DENV) outbreaks has traditionally relied on virus isolation and sequencing of the envelope gene [11, 13, 14]. Envelope gene sequencing remains an essential tool, providing the phylogeographic framework needed to compare local outbreaks with decades of global surveillance data. More recently, direct whole genome sequencing (WGS) of patient serum samples has enabled higher-resolution insights into viral transmission, lineage turnover, and regional movement of DENV [14–18]. Despite these advances, application of WGS for surveillance of outbreaks and endemic circulation in Pacific Island countries has remained limited, reflecting broader challenges around sustainable investment and equitable access to pathogen genomics in the region as highlighted by a recent commentary in *The Lancet Regional Health – Western Pacific* [19]. Moreover, this lack of genome representation has limited our understanding of the epidemiology and transmission pathways of dengue virus in the Pacific Islands. Here, we reconstruct the genomic epidemiology of DENV during the recent activity period (2023–2025) in Solomon Islands. Our objectives were to (1) generate whole genome sequences directly from patient serum samples, including those with low viral titres, (2) characterise the genomic epidemiology of circulating strains, and (3) investigate endemic circulation and outbreak resurgence events.

## Results

We first collected data on rapid diagnostic testing (RDT) from the National Referral Hospital (NRH) in Honiara (Fig. 1A, B), including gaps from national stock shortages of diagnostic tests) and nationally available reporting on dengue like illness (DLI) from sentinel sites (Fig. 1A, C, Table D1). We note an increase in DLI reported after the cessation of travel restrictions in July 2022 (Fig. 1C) prompting concerns about a delayed outbreak and initiation of the enhanced surveillance program. Despite the small sample size and gaps in referral for WGS due to RDT shortages, there were strong indications for a serotype changeover captured during the study period, as indicated by PCR typing and confirmed by WGS (Fig. 1D, E). In 2023, the vast majority of the strains were DENV-2 (n = 36) with single detection of DENV-4. This was contrasted by a single DENV-2 strain detected in 2024 (among the lower DLI incidence rates, Fig. 1C) and the majority of strains identified as DENV-1 (n = 7) or DENV-4 (n = 11). Previously a single whole genome sequence for the DENV-2 outbreak strain from 2016-2017 had been sequenced (NCBI:MH985858) which was derived from a Solomon Islands resident presenting in Queensland, Australia [20]. No other whole genome sequences from Solomon Islands exist. Our enhanced surveillance program therefore targeted acute cases confirmed by serology (NS1+) and serotype PCR at NRH and VIDRL collected between 2023 - 2025 (Fig. 1D, E).

**Figure 1.**
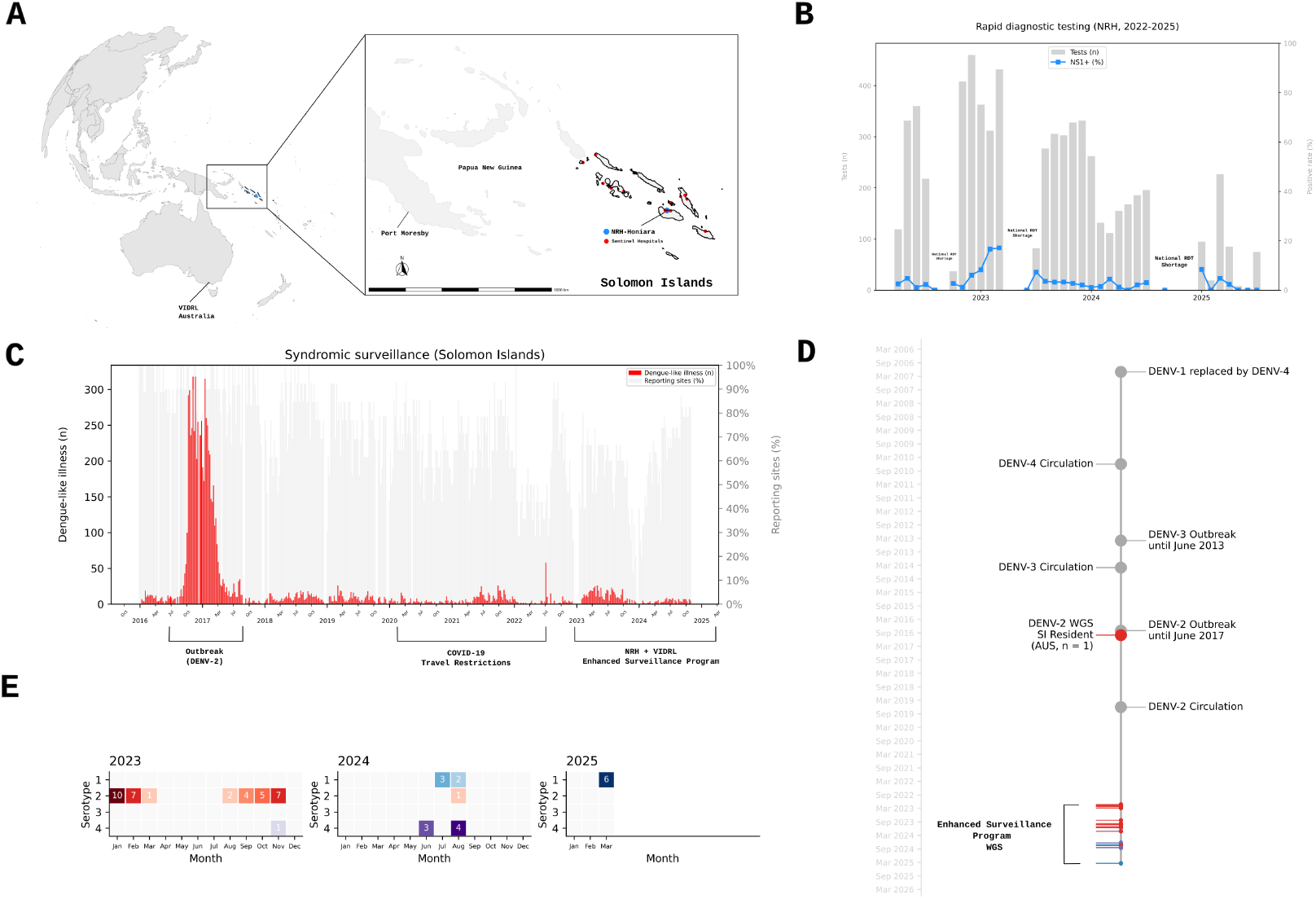
Dengue virus (DENV) outbreaks and whole genome sequencing in Solomon Islands. **(A)** Map displaying neighbouring Papua New Guinea and Solomon Islands Dengue Like Illness (DLI) surveillance reporting sites for syndromic and rapid serological DLI testing (NS1 + IgM + IgG). Sentinel hospitals and clinics (red circles) and the tertiary National Referral Hospital (NRH) in Honiara (blue circle). **(B)** NRH testing of DLI presenting cases and confirmed Dengue infection rates (NS1+) for 2023-2025. Reagent shortages of Rapid Diagnostic Tests (RDTs) at the NRH occurred at multiple points during the study timeline. **(C)** National Syndromic DLI reporting numbers for 2016-2025 (red) with reporting sites (grey). **(D)** Timeline of confirmed DENV outbreaks and serotype turnovers reported for Solomon Islands between 2013 and 2025. Red (DENV-2) and blue (DENV-1, DENV-4) lines indicate whole genomes recovered to date, with the only other whole genome DENV sequence before this study generated from a resident of Solomon Islands in Australia (DENV-2, 2016). **(E)** Serotype, sampling date and sample number of NRH DENV NS1+ referred samples received at VIDRL (excluding samples without recorded collection date, n = 5).

In total, 63 of the 249 confirmed NS1+ NRH cases were referred for WGS during this time frame (Fig. 2A) with around 20% of each serotype confirmed by NRH failing PCR confirmation at VIDRL and an overall agreement of 76.2% (100% excluding failing confirmatory PCRs) without off-target calls across serotypes (Fig. 2B). Of the 50 samples confirmed positive by VIDRL (79.4%) [21], 90% (45/50) were successfully sequenced (defined as average depth to respective serotype reference genomes ≥ 10x and consensus genome ≥ 60% genome coverage) (Fig. 2A). Two samples failed sequencing (Ct 38-40), while genome coverage for the remaining three ranged from 41.8% to 59.5% (Ct 28-35). Average genome coverage for respective DENV genotypes was 88.6% (DENV-1), 91.9% (DENV-2) and 82.0% (DENV-4). Regression analysis of cycle thresholds and genome coverage for pan-DENV and serotype-DENV PCRs indicated increased genome coverage with higher viral titres (Fig. 2C). Average age of NRH cases with a successful genome recovery (29.6 years) was representative of Solomon Islands national average age of acutely infected (NS1+) DLI patients (27.7 years). IgG seropositivity was also assessed: 47.6% of all cases (n = 30) showed evidence of prior DENV exposure (IgG-positive), while 30.2% (n = 19) were IgG-negative, indicating a primary DENV infection. IgG status was unknown for 22% of cases (n = 14, Table D1).

**Figure 2.**
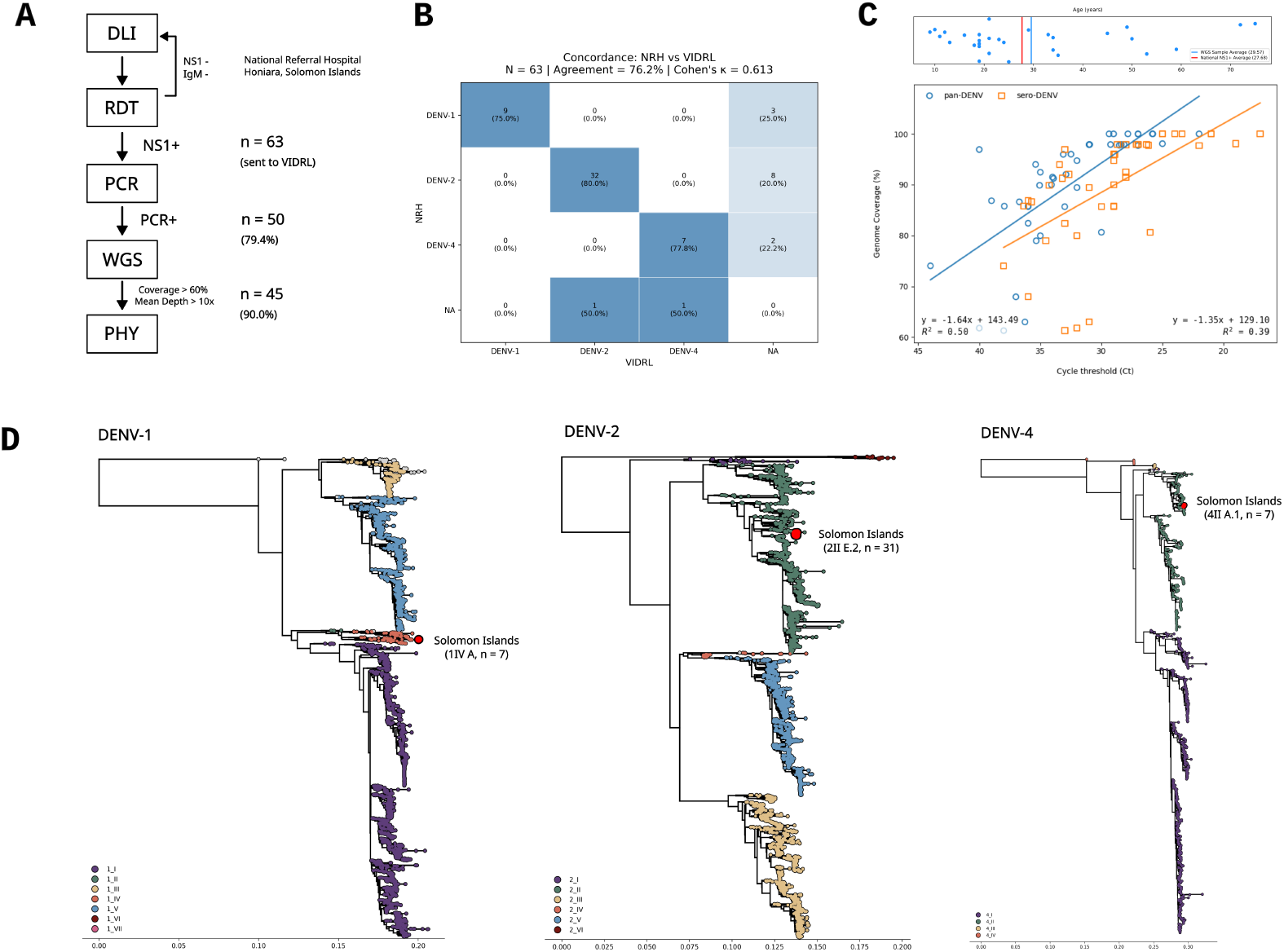
Dengue virus (DENV) testing and genome recovery in Solomon Islands and Australia. **(A)** Identification of samples used for genome recovery attempts based on acute infections (NS1+) detected at the National Referral Hospital (NRH) in Honiara and PCR confirmation after shipment to the Victorian Infectious Diseases Reference Laboratory (VIDRL) in Australia. **(B)** Agreement of serotype PCR at NRH and confirmatory serotype PCR at VIDRL. Disagreement between serotype-positive tests in Solomon Islands and failed confirmatory tests (NA) in Australia was likely due to time to shipment, sample volume, storage conditions and nucleic acid degradation. Confirmatory tests that passed quality control validate the serotype testing at NRH (100%, excluding NA). **(C)** Correlation of PCR cycle threshold (Ct) values with percentage of genome coverage (> 60%) from consensus genomes produced with the direct amplicon sequencing protocol. Age distribution of positive cases in this study (blue) and national average age of positive cases in Solomon Islands (red) is shown in the plot above. **(D)** Reconstruction of the dengue lineages whole genome dataset as serotype specific background phylogenies with the genomes recovered in this study in red with annotations (DENV-1, DENV-2, DENV-4).

Whole genome analysis confirmed PCR serotyping results (100% concordance, Table D1) for serotype 1 genotype IV lineage A (n = 7), serotype 2 genotype II lineage E.2 (n = 31) and serotype 4 genotype II lineage A.1 (n = 7) [21]. Placement within the reduced lineage-typing background phylogenies indicated three distinct clades for each serotype respectively (Fig. 2D). To better understand the geographical and temporal context of these clades, we reconstructed whole genome phylogenies and additionally combined them with broader envelope gene surveillance data available in public archives (August 2025). WGS phylogenies reconstructed the same singular clades as observed in the lineage typing background (Fig. 3, Fig. 4).

**Figure 3.**
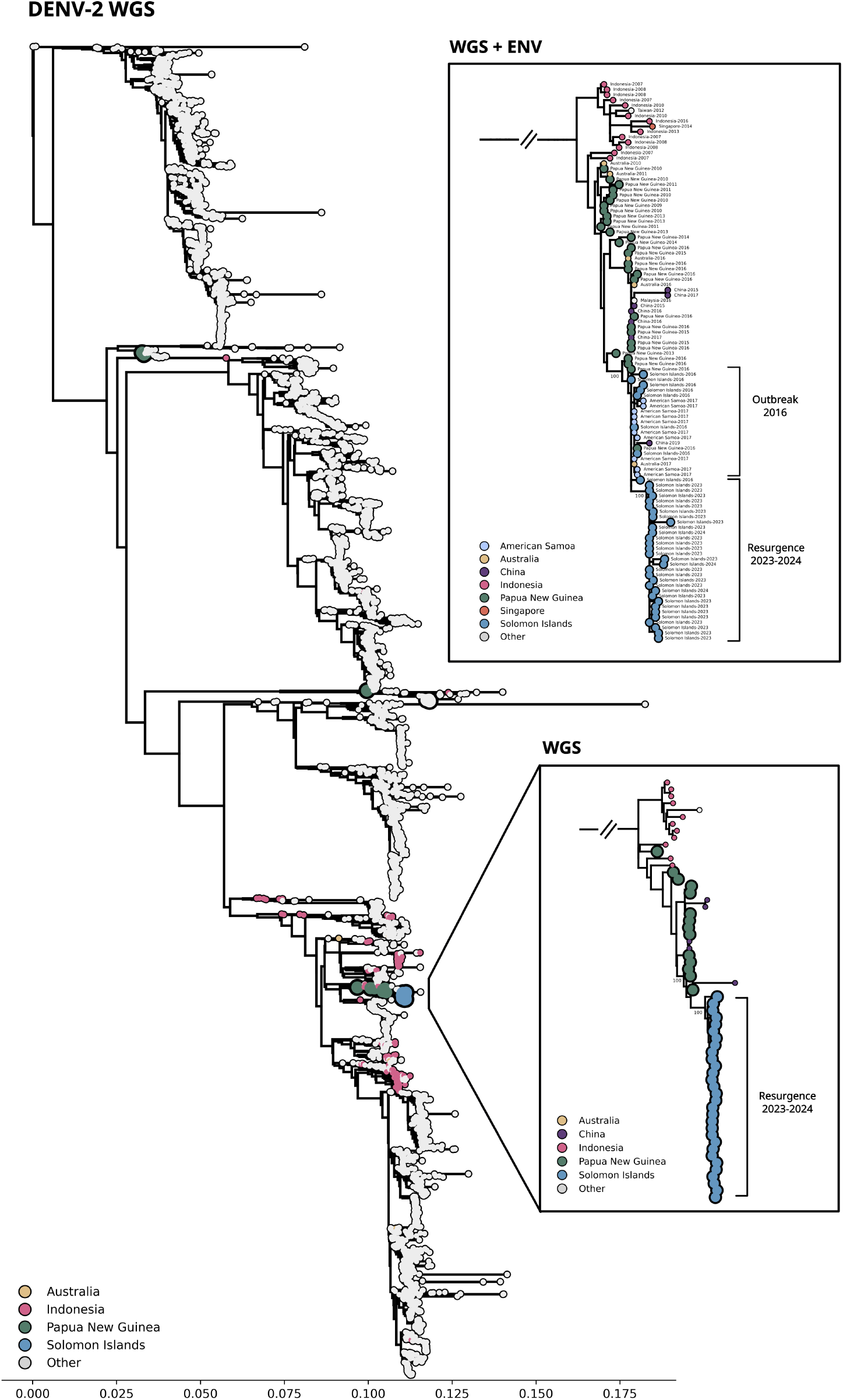
Dengue virus (DENV-2) global maximum likelihood phylogeny based on publicly available whole genome sequences (WGS). Insets show subtrees with the immediate geographical and temporal (annotations) context of Solomon Islands strains (blue) from WGS alone and combined with envelope gene (WGS + ENV) surveillance data from NCBI. One strain with excessive branch length was removed from the envelope gene subtree (VIDRL-DN0051).

**Figure 4.**
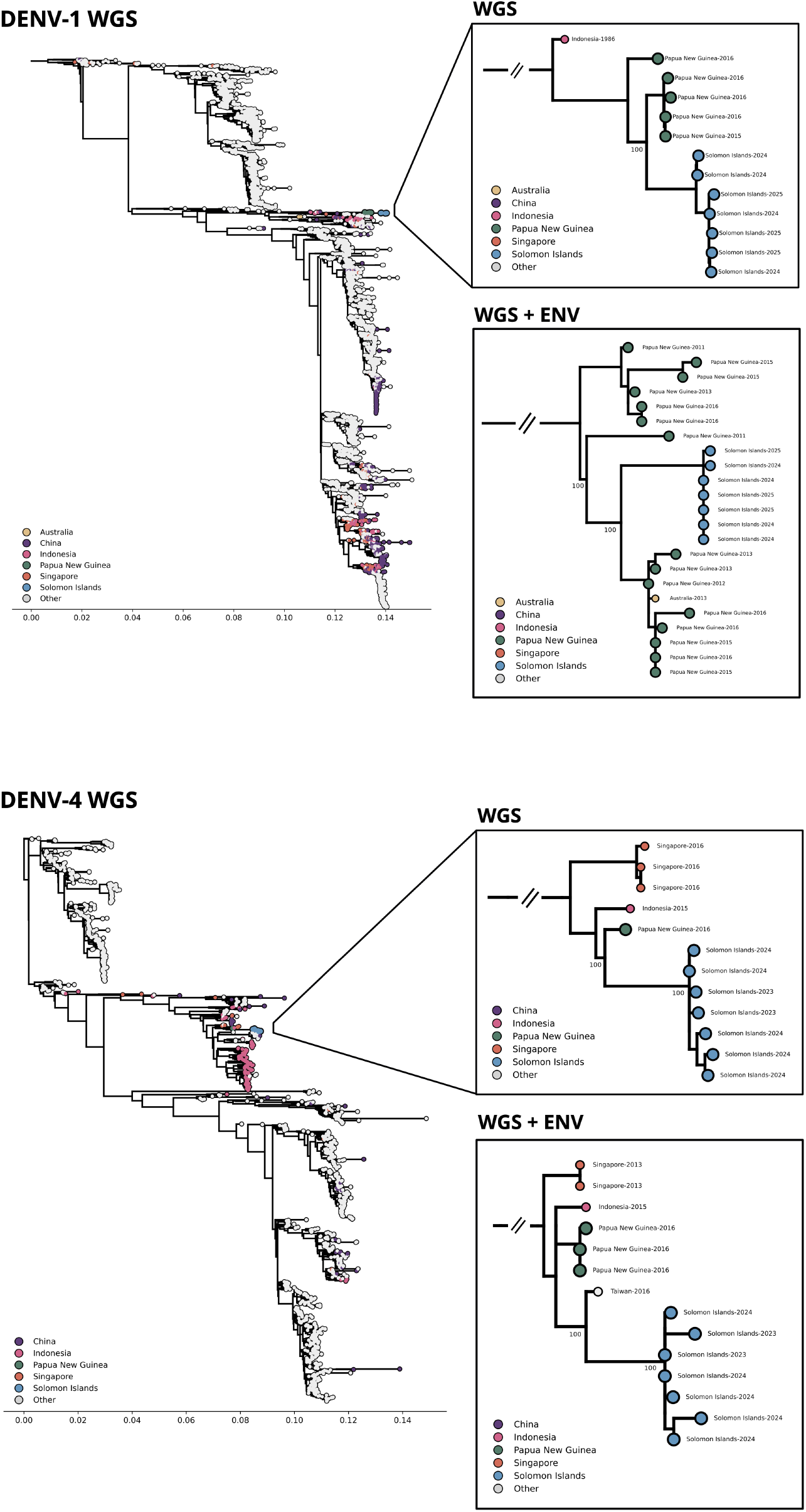
Dengue virus (DENV-1 and DENV-4) global maximum likelihood phylogeny based on publicly available whole genome sequences (WGS). Insets show subtrees the immediate geographical and temporal (annotations) of Solomon Islands strains (blue) from WGS alone and combined with envelope gene (WGS + ENV) surveillance data from NCBI.

Envelope gene data was essential to contextualize the DENV-2 strains sampled in 2023-2024 as a continuation of the outbreak strain circulating in Solomon Islands since 2016 (Fig. 3, insets). WGS and envelope gene data strongly suggest that this strain originated from Papua New Guinea. Similarly, WGS and envelope gene data demonstrated that the DENV-1 Solomon Island clade was nested among strains from Papua New Guinea (Fig. 4, insets). While there were indications for this association for DENV-4 in the WGS data, additional envelope gene data identified a strain from Taiwan (2016) as the closest related strain to the clade, suggesting that more extensive sampling in the Western Pacific region may uncover previously hidden patterns of transmission across the region, and reduce the impact of sampling bias on phylogeographic inference from limited public data (Fig. 4, insets).

## Discussion

In this study, we performed WGS of 50 NS1+, PCR-confirmed DENV serum samples collected at the National Referral Hospital (NRH) in Solomon Islands to evaluate the genomic epidemiology of DENV during the post–COVID-19 resurgence period (2023–2025). These genomes represent the first such data from the country and the largest set of primary sample whole genomes from a Pacific Island country to date [14–16]. Sequencing was conducted through referral to a partner laboratory (VIDRL) in Melbourne, Australia, highlighting the value of regional collaboration in bridging immediate capacity gaps. PCR and WGS at VIDRL validated the DLI screening methods and molecular serotype testing at NRH with high concordance (100%, excluding failed confirmatory PCRs) showcasing the reliability of local diagnostic screening in Solomon Islands. Whole genome recovery using amplicon schemes [22] directly from acute infections (NS1+) serum samples was highly effective, with a 90% recovery rate and >80% average genome coverage across DENV serotypes, including successful reconstruction from low-titre samples (Ct 38–44). Resulting genomes were suitable for detailed lineage typing [21] and phylogenetic reconstruction, providing the resolution needed to investigate local and regional transmission dynamics alongside existing envelope gene surveillance from the Western Pacific region for broader geographical contextualisation. These findings provide evidence of high-quality genome recovery under real-world conditions and highlight the value of integrated diagnostic and sequencing workflows across institutional partnerships.

Previous genomic DENV studies in Pacific Island countries and territories have been limited in their sampling scope and genome recovery success. Descloux and colleagues [15] used an early amplicon scheme to generate 12 near-complete DENV-1 genomes from endemic circulation in French Polynesia between 2001 and 2006, though without systematic reporting of recovery rates. Inizan and colleagues [23] employed metagenomic sequencing of patient samples during two DENV-1 outbreaks in New Caledonia (2013–2016), achieving genome recovery rates of 17.6% (3/17) and 30.4% (21/69). Jonduo and colleagues [16] generated 10 DENV-1 and DENV-2 whole genomes from Papua New Guinea using metagenomic sequencing, but recovery was restricted to samples with low cycle thresholds (Ct < 28). In contrast, high recovery rates and genome coverage from a wide range of viral titres were achieved in this study, demonstrating the technical feasibility of applying amplicon-based sequencing protocols [22] to a broader range of patient sera. This represents a significant change in the quality and consistency of dengue genomic data available from Pacific Island countries and territories, enabling lineage typing [21] and robust phylogenetic contextualisation of local outbreaks within the wider regional transmission network. As routine data generation becomes feasible whole genome sequences will allow for the investigation of local transmission pathways and integration with genomic vector-borne disease surveillance.

The resurgence and serotype turnover observed in Solomon Islands during this study highlights a common pattern across Pacific Island countries, where non-endemic serotypes are introduced into the population, circulate transiently, and subsequently undergo replacement events [4]. Our data show a shift from predominance of DENV-2 in 2023 to increased circulation of DENV-1 and DENV-4 in 2024, consistent with epidemiological models where high seroprevalence and population immunity shape outbreak dynamics [4, 11]. Co-circulation of multiple serotypes raises the risk of severe disease and complicates outbreak response, reinforcing the value of local diagnostics and genome sequencing to monitor introductions and turnover events. Phylogenetic analyses indicate that the 2023–2024 Solomon Islands DENV-2 clade, as well as the emerging DENV-1 clade, are closely related to strains circulating in Papua New Guinea. These findings support a model of continued cross-border viral circulation between Papua New Guinea and Solomon Islands, consistent with earlier hypotheses that Papua New Guinea may act as a “stepping stone” for dissemination into neighbouring Pacific Island countries and territories [16, 24]. While WGS data suggested possible links to Papua New Guinea for DENV-4, envelope gene comparisons instead identified a strain from Taiwan (2016) as the nearest neighbour. This highlights the importance of broader sampling across the Western Pacific Region, where gaps in local and global sequence availability may obscure true transmission pathways. Limited numbers of genomes and sparse regional sampling mean that unsampled diversity, including additional lineages of DENV-1 and DENV-4, cannot be ruled out. Leveraging from existing genomic surveillance programme from WHO or CoViNet can help expand DENV surveillance programs across the Pacific Island countries and territories, critical in reducing bias and accurately reconstructing viral movement in the region. Understanding the DENV transmission network prior to the introduction of new DENV serotypes is important in informing effective public health control strategies across the whole Western Pacific regions, with supporting phylogenetic evidence presented in this study.

Our study has several limitations. First, genomic comparisons are constrained by a lack of historical Solomon Islands data, with the 2016 outbreak represented only by envelope gene sequences and a single DENV-2 genome sequenced from a resident presenting in Australia [20]. In addition, national shortages in the availability of rapid diagnostic testing units caused gaps in surveillance during the study period. Second, while referral sequencing in Australia provided proof-of-concept, local sequencing capacity remains absent, limiting timeliness of genomic data for outbreak response. Third, the lack of linked clinical and hospitalisation data restricted our ability to correlate genomic findings with disease severity and patient outcomes. These limitations reflect broader systemic reporting barriers in routine genomic surveillance across the Pacific [4].

Outcomes from this study echo recent calls from *The Lancet Regional Health – Western Pacific* [19] for region-specific evidence to inform the integration of pathogen genomics into public health practice. As emphasised in the commentary, scaling up genomic surveillance in the Western Pacific requires technical demonstration and contextual evaluation. By providing practical evidence on genome recovery rates, completeness, and downstream analytical utility, this study contributes towards the regional evidence base needed to guide investment in sequencing infrastructure and assess public health benefit. Furthermore, by establishing proof-of-concept for high-quality DENV sequencing from primary patient samples through referral partnerships with Australia, this work lays a foundation for Solomon Islands to progressively build local sequencing capacity. In this step wise model, leveraging external infrastructure while generating locally relevant data aligns with the broader regional need for sustainable and equitable implementation of pathogen genomics beyond high-income settings. Ongoing regional DENV outbreaks in 2025 with thousands of confirmed cases across Pacific Island countries and territories including Fiji, American Samoa, Kiribati, Nauru and Tuvalu [25], reinforce the urgency of embedding genomic tools into Pacific surveillance systems to ensure equitable access to actionable data across the region.

In conclusion, this study demonstrates the feasibility and utility of WGS for dengue surveillance in Solomon Islands, while exposing critical gaps in regional genomic representation of DENV. By leveraging referral sequencing as a transitional model, we provide an example for the initial stages of local capacity-building in Solomon Islands and neighbouring Pacific Island countries and territories. Our study provides the evidence that strengthening DENV genomic surveillance in the pacific island countries is of broader geographical interest and will improve outbreak detection and support timely public health responses in the context of climate-driven changes to arboviral transmission.

## Methods

### Ethics and data sharing

This study is approved by the Solomon Island MOHMS. Local ethical approval was granted in 2026 by the Research Ethics Committee, Solomon Islands Health Research and Ethics Review Board (SIHRERB, HRE024/25) and at Melbourne Health (QA2024127).

### Study design

This study was conceived as part of an enhanced surveillance program to monitor the circulating DENV serotype(s) via DLI cases using molecular characterisation and whole genome sequencing (WGS). The scope of this study did not contain any vector-based components including mosquito surveillance or vector sequencing. This study spanned the surveillance period of January 2023 to March 2025 and involved individuals presenting to emergency departments with DLI manifestations at the Solomon Islands National Referral Hospital (NRH) and sentinel testing clinics located in the capital (Honiara). Sequencing cohort selection was based on both patient and laboratory characteristics (Fig S1), including (1) presentation to hospital or clinics with DLI (2) positive test from rapid diagnostic tests (RDT) or PCR at NRH (3) availability of RDT and PCR reagents as initial screening test (4) adequate sample volume (≥200 *μ*l) for VIDRL referral testing and WGS. NRH referred five shipments of laboratory-confirmed NS1 real time positive DENV blood specimens (n = 63) to VIDRL for confirmatory referral testing and next-generation sequencing (Fig. S1)). Detailed clinical and epidemiological data was not available. Where available, additional demographic and laboratory testing details (age, gender, DLI presenting location, NS1/IgG/IgM status) of referred samples were collated retrospectively by NRH staff (Table D1).

### Dengue case definitions

Solomon Islands and other PICs report DENV cases to the Pacific Syndromic Surveillance System (PSSS) in accordance with the World Health Organization (WHO) 2009 Dengue Classification scheme [[26]]. A suspected case (Dengue-Like Illness, DLI) is defined as a healthcare seeking individual with an acute febrile illness (≥ 38 °C for ≥ 2 days) with ≥ 2 dengue fever symptoms (nausea, vomiting, myalgia, macular or maculopapular rash, headache and/or retro-orbital pain) who recently resided in or travel within a dengue-endemic area ≤ 14 days and a negative Malaria test (RDT or microscopy parasite smear). A confirmed case of acute DENV infection requires both clinical presentation and diagnostic laboratory evidence. Laboratory confirmation methods include the direct detection of antigen non-structural protein 1 (NS1) and/or anti-DENV immunoglobulin M antibodies (IgM) via Enzyme-Linked Immunosorbent Assay (ELISA) and/or Rapid Diagnostic Tests (RDT), and/or presence of DENV RNA by real-time reverse transcription polymerase chain reaction (RT-PCR) in serum.

### Diagnostic molecular testing and screening

At the National Referral Hospital (NRH), DENV infection status in patients presenting with DLI is assessed using the Abbott Bioline™ Dengue Duo RDT, which detects for the presence or absence of NS1 antigen and anti-DENV IgG/IgM antibodies. NS1-positive blood samples undergo further testing, beginning with nucleic acid extraction using the Qiagen QIAamp Viral RNA Mini Kit. DENV molecular and serotype characterization is performed using the CDC Trioplex real-time PCR assay (DNature Trioplex) with the Invitrogen SuperScript™ III Platinum One-Step RT-PCR Kit on the ABI 7500 Fast system following manufacturer’s instructions. For NGS at the reference laboratory (VIDRL), blood samples were extracted using MagMAX™ Viral/Pathogen II (MVP II) Nucleic Acid Isolation Kit according to the manufacturer (Thermofisher, Australia). Prior to NGS, all samples underwent in-house real-time PCR methods (pan and serotype specific DENV PCR) using standard ABI 7500 fast default cycling conditions, via two-step cycling using BIOLINE cDNA Synthesis Kit and SensiFAST™ Probe Lo-ROX real time PCR Kit. Primer/probe sequences and cycling conditions are available on request.

### Direct whole genome amplicon sequencing

Irrespective of Ct values (17-44), direct sequencing of DENV was attempted on all real-time PCR positive samples (n=50) using the sero-specific primer schemes (DENV-1, DENV-2, DENV-4) published for the ‘DengueSeq’ V2 & V3 protocols [22]. Libraries were generated using the Illumina COVIDSeq Test Kit (RUO kit) with IDT for Illumina Nextera DNA Unique Dual Indexes Sets A-D. Final library was diluted to 75pM and included a 1% PhiX control with sequencing on Illumina iSeq100 platform and i1 Flow Cell with 150 bp paired-end reads and adapter trimming (Illumina, Australia). Read files were processed with the associated DengueSeq pipeline [22] with default parameters (https://github.com/grubaughlab/DENV_pipeline). Successful genome recovery was defined as consensus genomes ≥ 10x average depth and ≥ 60% genome coverage.

### Lineage scheme typing and phylogenies

Genotypes and lineages were assigned using the newly WHO endorsed dengue lineages scheme [21] with Nextclade v3.16 (https://clades.nextstrain.org/) [27]. Curated background genomes (2024-03-29) of dengue lineages for phylogenetic reconstruction of each serotype were downloaded (https://github.com/grubaughlab/DENV-genomics) and combined with the genomes sequenced in this study. Augur v26.0.0 [28] alignment and tree building commands with default configuration and the HKY model were used to construct the lineage phylogenies (Fig. 2D). Traditional bootstrap values (-b 1000) of divergences relevant to interpretation are annotated in the figures.

### Data collection for phylogeography

Broad geographic coverage and accurate annotation of geographic origin are important to minimize bias in interpretation of regional transmission pathways and phylogeographic models when using sequence data from public archives. We first collected contextual whole genome (WGS) and envelope gene (E-gene) sequences for the serotypes detected in Solomon Islands (DENV-1, DENV-2 and DENV-4) from NCBI Virus (August 2025) with a sequence length filter between 10000 bp and 12000 bp for complete genomes and between 1000 bp and 3000 bp for putative envelope gene sequences. We removed all sequences without collection date or geographic location (country of origin) and filtered sequences with completeness < 70% and > 5% ambiguous nucleotides. We then added Solomon Islands whole genome sequences with > 60% completeness (n = 45) generated in this study to the WGS dataset (*n*_*DENV* −1_ = 6189, *n*_*DENV* −2_ = 5511, *n*_*DENV* −4_ = 1143). Finally we added the putative envelope gene sequences for a combined WGS dataset (*n*_*DENV* −1_ = 15391, *n*_*DENV* −2_ = 11678, *n*_*DENV* −4_ = 3226). Sentinel sites reporting DLI across provinces (Table S1) were mapped using R packages rnaturalearth v1.0.1 (https://github.com/ropensci/rnaturalearthhires) and ggspatial v1.1.9 (https://github.com/paleolimbot/ggspatial) (Fig. 1A).

### Maximum likelihood phylogenies

Genomes were aligned against their respective serotype reference genome (DENV-1: NC_001477.1, DENV-2: NC_001474.2, DENV-4: NC_002640.1) with MAFFT v.7.490 [29] as part of the Augur v26.0.0 alignment pipeline. Reference genomes were excluded from the alignment and gaps filled with missing characters (N). We used the locations of the envelope gene of the reference genome annotation to subset the combined sequence alignment to derive envelope gene phylogenies for each serotype and filtered any sequences from the alignment with completeness < 60%. Maximum-likelihood phylogenies were constructed using IQTree v2.1.4 [30] and the HKY model in the Augur tree building pipeline for both WGS and envelope gene alignments. We plotted root-to-tip distance regressions against the sampling date and computed z-scores from residuals to automatically exclude sequences with outlier branch lengths above a serotype specific z-score threshold (*n*_*DENV* −1_ > 3.0, *n*_*DENV* −2_ > 3.0, *n*_*DENV* −4_ > 2.0) using viremic v0.1.0 (https://github.com/esteinig/viremic). Trees were visualized with custom Python scripts using ETE3 toolkit v4.4.0 [31] and the baltic library v0.3.0 (https://github.com/evogytis/baltic).

## Data availability

Genome sequences are available at NCBI (Table D1).

## Acknowledgements

The authors would like to acknowledge and thank laboratory staff from the National Referral Hospital and Solomon Islands Ministry of Health and Medical Services for coordinating sample referrals. The authors are grateful for the in-kind sequencing and testing support provided by the VIDRL Translational Diagnostics and Viral Identification laboratories.

## Data availability

Consensus genome sequences are available at NCBI under accessions listed in Table D1.

## Author Contributions

| Contribution | Conceptualization | Methodology | Software | Validation | Formal Analysis | Investigation | Data Curation | Resources | Writing - Original Draft | Writing - Review & Editing | Visualization | Supervision | Project Administration | Funding Acquisition |
| --- | --- | --- | --- | --- | --- | --- | --- | --- | --- | --- | --- | --- | --- | --- |
| Jean Moselen | X | X |  |  | X | X | X |  | X | X | X | X | X |  |
| Eike Steinig | X | X | X | X | X | X | X |  | X | X | X | X |  |  |
| Andrew Darcy | X |  |  |  |  | X | X | X |  | X |  |  |  |  |
| Alfred Dofai | X |  |  |  |  | X | X | X |  | X |  |  |  |  |
| Angella Manele |  |  |  |  |  | X | X | X |  | X |  |  |  |  |
| Brenda Lauri |  |  |  |  |  | X |  | X |  | X |  |  |  |  |
| Cynthia Joshua |  |  |  |  |  | X | X | X |  | X |  |  |  |  |
| Paul Mauruwai |  |  |  |  |  | X |  | X |  | X |  |  |  |  |
| Ammar Aziz |  | X | X |  | X |  |  |  |  | X |  |  |  |  |
| Nicole Orlando |  | X |  |  |  | X | X |  |  | X |  |  |  |  |
| Paul Horwood |  |  |  |  |  |  |  |  |  | X |  |  |  |  |
| Leon Caly |  |  |  |  |  |  |  | X |  | X |  |  |  |  |
| Navin Karan |  |  |  |  |  |  | X | X |  | X | X |  | X |  |
| Janella Solomon | X |  |  |  |  |  |  | X |  | X |  | X | X | X |
| Chuan K. Lim | X |  |  |  |  |  |  | X |  | X | X | X | X | X |

## Supplementary Information

**Table S1.** Hospitals and medical centres in the Solomon Islands sentinel surveillance network with activity status reporting dengue-like illness in 2023 and 2024.

| Name | Facility | Province | Region | Island | 2023 | 2024 |
| --- | --- | --- | --- | --- | --- | --- |
| Atoifi Adventist Hospital | Church Hospital | Malaita | Eastern | Malaita | Yes | No |
| Auki Dispensary | Urban Health Centre | Malaita | Central | Malaita | Yes | Yes |
| Gizo Hospital | Provincial Hospital | Western | – | Ghizo Island | Yes | Yes |
| Helena Goldie Hospital | Church Hospital | Western | – | New Georgia | Yes | Yes |
| Kilu'ufi Hospital | Provincial Hospital | Malaita | Central | Malaita | Yes | Yes |
| KiraKira Hospital | Provincial Hospital | Makira-Ulawa | – | Makira (San Cristobal) | Yes | Yes |
| Kukum Clinic | Area Health Centre | Guadalcanal | Honiara | Guadalcanal | Yes | No |
| Lata Hospital | Provincial Hospital | Temotu | – | Nendo | Yes | Yes |
| Mataniko Health Centre | Area Health Centre | Guadalcanal | Honiara | Guadalcanal | Yes | Yes |
| National Referral Hospital | Tertiary Hospital | Guadalcanal | Honiara | Guadalcanal | Yes | Yes |
| Rove Area Health Centre | Area Health Centre | Guadalcanal | Honiara | Guadalcanal | Yes | Yes |
| Taro Hospital | Provincial Hospital | Choiseul | – | Taro | Yes | Yes |
| Good Samaritan Hospital | Church Hospital | Guadalcanal | – | Guadalcanal | No | Yes |
| Mbokona Urban Health Clinic | Rural Health Centre | Guadalcanal | Honiara | Guadalcanal | No | Yes |
| Mbokonaavera Urban Clinic | Rural Health Centre | Guadalcanal | Honiara | Guadalcanal | No | Yes |
| Naha Urban Clinic | Rural Health Centre | Guadalcanal | Honiara | Guadalcanal | No | Yes |
| Nila Area Health Centre | Area Health Centre | Western | – | Poporang | No | Yes |
| Seghe Area Health Centre | Area Health Centre | Western | – | New Georgia | No | Yes |
| Tulagi Provincial Hospital | Provincial Hospital | Central | – | Nggela Sule | No | Yes |
| Vura Urban Clinic | Rural Health Centre | Guadalcanal | Honiara | Guadalcanal | No | Yes |
| White River Health Clinic | Rural Health Centre | Guadalcanal | Honiara | Guadalcanal | No | Yes |

**Figure S1.**
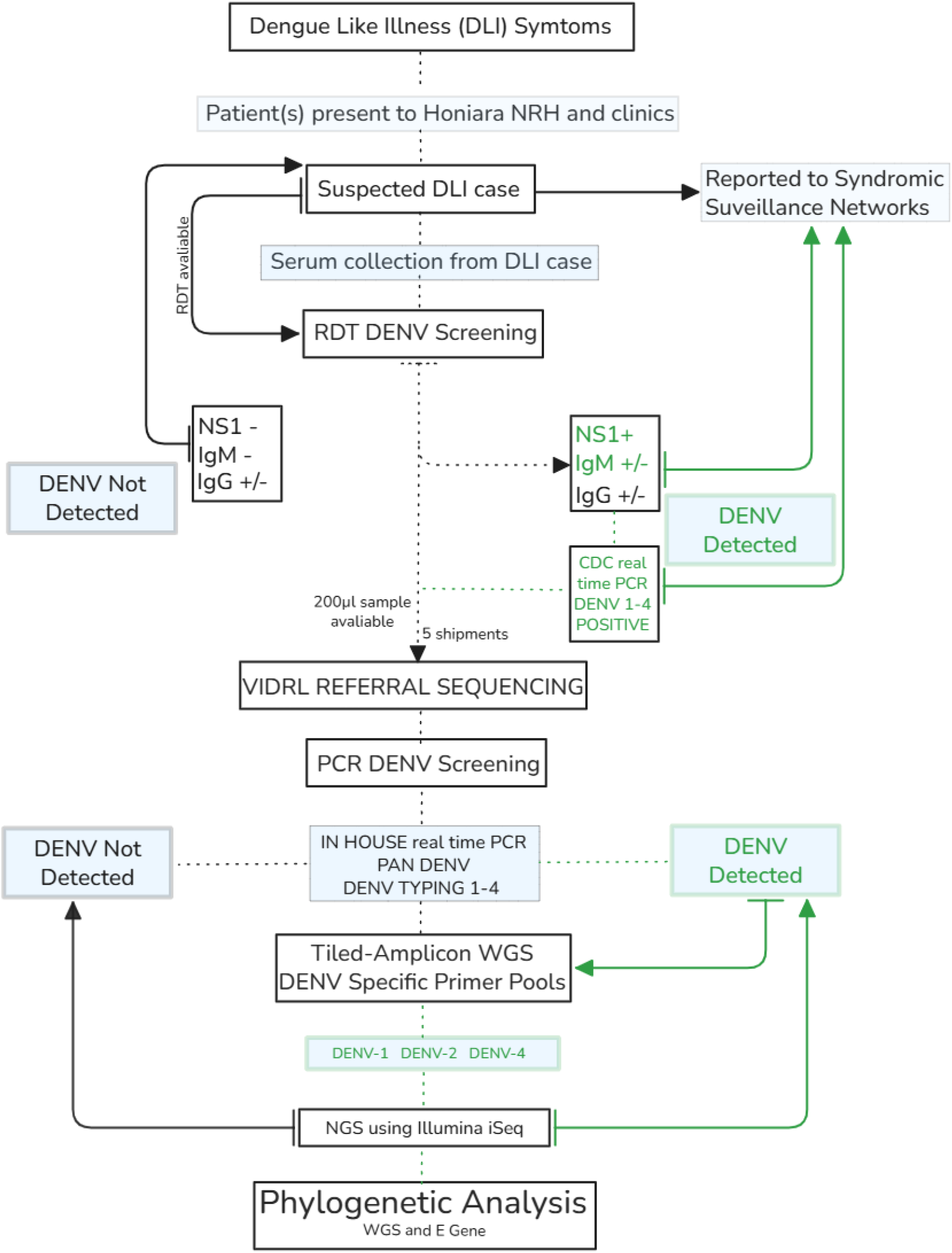
Study design and diagnostic referral workflow for the National Referral Hospital in Honiara, Solomon Islands and the Victorian Infectious Diseases Reference Laboratory (VIDRL) in Melbourne, Australia.

## Appendix 1

### Research in context

#### Evidence before this study

Dengue virus (DENV) is hyper-endemic across the Western Pacific region, with frequent outbreaks and documented serotype displacement in many Pacific Island countries and territories. While whole genome sequencing (WGS) of DENV from primary samples has increasingly been applied in Southeast Asia and other higher-resource settings, its use in Pacific Island countries has been sparse. We conducted a PubMed search until August 2025 with the search terms “pacific islands”, “dengue”, “whole genome sequencing” to evaluate studies using direct-from-sample whole genome sequencing of dengue virus in the Pacific Islands. We additionally searched for available complete genome sequences on NCBI Virus with taxonomic qualifiers “Dengue virus”, “Orthoflavivirus denguei”, global and regional setting “Oceania” and minimum sequence length of 8000 bp. Published studies from the region using whole genome from primary samples (n = 3) have typically involved small numbers of genomes (n = 10 – 24), limited recovery success (17.6% - 30.4%), or metagenomic approaches restricted to high viral titre samples (Ct < 28). Prior to this study, no whole genome dengue virus sequences had been generated directly from patient serum samples collected in the Solomon Islands, and genomic data from Pacific Island countries remained under-represented on NCBI Virus (169 out of 19699 genomes were from Pacific Island countries). This lack of genomic representation has constrained understanding of dengue transmission pathways, serotype turnover, and regional connectivity, and has limited the evidence base needed to support equitable implementation of pathogen genomics in Pacific public health systems.

#### Added value of this study

This study reports the first whole genome dengue virus sequences generated from patient serum samples collected in the Solomon Islands and represents the largest set of primary-sample dengue genomes produced from a Pacific Island country to date. Using an amplicon-based sequencing approach and a referral laboratory model, we demonstrate high genome recovery rates across a wide range of viral titres, including low-titre samples, under real-world surveillance conditions. Genomic analysis confirms co-circulation of DENV-1, DENV-2, and DENV-4 during the post–COVID-19 period, with evidence of serotype turnover emerging in 2024. Phylogeographic analyses place circulating Solomon Islands strains within regional transmission networks, with strong support for ancestral introductions from Papua New Guinea for DENV-1 and DENV-2.

#### Implications of all the available evidence

In combination with existing epidemiological and molecular data, our findings show that whole genome sequencing of dengue virus from primary sample is technically feasible, epidemiologically informative and operationally valuable for surveillance in Pacific Island settings. Referral-based sequencing partnerships can provide an effective transitional model for generating high-quality genomic data while local molecular testing capacity is validated and progressively developed. Expanding dengue genomic surveillance across Pacific Island countries and territories will improve detection of serotype turnover, clarify regional transmission pathways, and support more timely and equitable public health responses. With critical gaps in DENV regional sampling and sequencing representation (and WHO reporting the highest dengue cases in the region since 2016) our genome-informed findings are a valuable contribution towards understanding the transmission and relatedness of DENV in the broader Pacific region.

**Table D1.** Summary of sample metadata and diagnostic results (NA = not available, ND = not detected).

| sample_id | genome_id | accession | shipment_date | collection_date | rdt_ns1 | rdt_igm | rdt_igg | nrh_serotype | vidrl_dengue_ct | vidrl_serotype_ct | pcr_serotype | wgs_serotype | wgs_coverage | wgs_lineage |
| --- | --- | --- | --- | --- | --- | --- | --- | --- | --- | --- | --- | --- | --- | --- |
| 01 | VIDRL-DN0001 | PX683040 | 2023-06 | 2023-01 | POS | POS | POS | 2 | 34 | 33 | DENV-2 | DENV-2 | 91.24 | 2II_E.2 |
| 02 | VIDRL-DN0002 | PX683041 | 2023-06 | 2023-02 | POS | POS | POS | 2 | 28 | 26 | DENV-2 | DENV-2 | 97.91 | 2II_E.2 |
| 03 | VIDRL-DN0003 | PX683042 | 2023-06 | 2023-01 | POS | POS | POS | 2 | 33 | 33 | DENV-2 | DENV-2 | 92.03 | 2II_E.2 |
| 04 | NA | NA | 2023-06 | 2023-03 | POS | POS | POS | 2 | ND | ND | NA | NA | NA | NA |
| 05 | VIDRL-DN0004 | PX683043 | 2023-06 | 2023-01 | POS | POS | POS | 2 | 35 | 33 | DENV-2 | DENV-2 | 93.98 | 2II_E.2 |
| 06 | VIDRL-DN0005 | PX683044 | 2023-06 | 2023-01 | POS | POS | NEG | 2 | 29 | 26 | DENV-2 | DENV-2 | 97.75 | 2II_E.2 |
| 07 | VIDRL-DN0006 | PX683045 | 2023-06 | 2023-01 | POS | POS | POS | 2 | 35 | 34 | DENV-2 | DENV-2 | 89.88 | 2II_E.2 |
| 08 | NA | NA | 2023-06 | 2023-01 | POS | POS | POS | 2 | 36 | ND | DENV | NA | NA | NA |
| 09 | VIDRL-DN0007 | PX683046 | 2023-06 | 2023-02 | POS | POS | POS | 2 | 29 | 27 | DENV-2 | DENV-2 | 97.91 | 2II_E.2 |
| 10 | VIDRL-DN0008 | PX683047 | 2023-06 | 2023-02 | POS | POS | POS | 2 | 38 | 36 | DENV-2 | DENV-2 | 85.78 | 2II_E.2 |
| 11 | NA | NA | 2023-06 | unknown | unknown | unknown | unknown | 2 | ND | ND | NA | NA | NA | NA |
| 12 | VIDRL-DN0009 | PX683048 | 2023-06 | 2023-01 | POS | POS | POS | 2 | 31 | 29 | DENV-2 | DENV-2 | 97.91 | 2II_E.2 |
| 13 | VIDRL-DN0010 | PX683049 | 2023-06 | 2023-02 | POS | POS | POS | 2 | 33 | 29 | DENV-2 | DENV-2 | 95.97 | 2II_E.2 |
| 14 | VIDRL-DN0011 | PX683050 | 2023-06 | 2023-01 | POS | POS | POS | 2 | 35 | 35 | DENV-2 | DENV-2 | 78.97 | 2II_E.2 |
| 15 | VIDRL-DN0012 | PX683051 | 2023-06 | 2023-01 | POS | POS | POS | 2 | 28 | 23 | DENV-2 | DENV-2 | 100 | 2II_E.2 |
| 16 | VIDRL-DN0013 | PX683052 | 2023-06 | 2023-01 | POS | POS | NEG | 2 | 28 | 27 | DENV-2 | DENV-2 | 97.91 | 2II_E.2 |
| 17 | NA | NA | 2023-06 | 2023-02 | unknown | unknown | unknown | 2 | ND | ND | NA | NA | NA | NA |
| 18 | VIDRL-DN0014 | PX683053 | 2023-06 | 2023-02 | unknown | unknown | unknown | 2 | 37 | 36 | DENV-2 | DENV-2 | 86.63 | 2II_E.2 |
| 19 | NA | NA | 2023-06 | 2023-02 | POS | POS | POS | 2 | ND | ND | NA | NA | NA | NA |
| 20 | VIDRL-DN0015 | PX683054 | 2023-11 | 2023-08 | POS | NEG | POS | 2 | 34 | 29 | DENV-2 | DENV-2 | 90 | 2II_E.2 |
| 21 | VIDRL-DN0016 | PX683055 | 2023-11 | 2023-08 | POS | NEG | POS | 2 | 32 | 29 | DENV-2 | DENV-2 | 96 | 2II_E.2 |
| 22 | NA | NA | 2023-11 | 2023-09 | POS | POS | POS | 2 | 34 | 40 | DENV-2 | NA | NA | NA |
| 23 | VIDRL-DN0017 | PX683056 | 2023-11 | 2023-09 | POS | POS | NEG | 2 | 26 | 21 | DENV-2 | DENV-2 | 100 | 2II_E.2 |
| 24 | VIDRL-DN0018 | PX683057 | 2023-11 | 2023-09 | POS | NEG | NEG | 2 | 26 | 21 | DENV-2 | DENV-2 | 100 | 2II_E.2 |
| 25 | NA | NA | 2023-11 | 2023-09 | NEG | NEG | NEG | 2 | ND | 38 | DENV-2 | NA | NA | NA |
| 26 | VIDRL-DN0019 | PX683058 | 2023-11 | 2023-10 | NEG | NEG | POS | 2 | 36 | 31 | DENV-2 | DENV-2 | 63 | 2II_E.2 |
| 27 | VIDRL-DN0020 | PX683059 | 2023-11 | 2023-10 | POS | NEG | NEG | 2 | 29 | 25 | DENV-2 | DENV-2 | 100 | 2II_E.2 |
| 28 | VIDRL-DN0021 | PX683060 | 2024-02 | 2023-10 | POS | POS | POS | 2 | 31 | 27 | DENV-2 | DENV-2 | 97.91 | 2II_E.2 |
| 29 | VIDRL-DN0022 | PX683061 | 2024-02 | 2023-11 | unknown | unknown | unknown | 2 | 32 | 29 | DENV-2 | DENV-2 | 94.73 | 2II_E.2 |
| 30 | VIDRL-DN0023 | PX683062 | 2024-02 | 2023-10 | POS | NEG | NEG | 2 | 31 | 28 | DENV-2 | DENV-2 | 97.87 | 2II_E.2 |
| 31 | NA | NA | 2024-02 | 2023-10 | POS | NEG | NEG | 2 | ND | ND | NA | NA | NA | NA |
| 32 | VIDRL-DN0024 | PX683083 | 2024-02 | 2023-11 | POS | NEG | POS | 2 | 37 | 36 | DENV-2 | DENV-2 | 67.94 | 2II_E.2 |
| 33 | NA | NA | 2024-02 | 2023-11 | POS | NEG | POS | 2 | ND | ND | NA | NA | NA | NA |
| 34 | NA | NA | 2024-02 | 2023-11 | POS | NEG | NEG | 2 | ND | ND | NA | NA | NA | NA |
| 35 | VIDRL-DN0025 | PX683063 | 2024-02 | 2023-11 | POS | NEG | NEG | 2 | 29 | 25 | DENV-2 | DENV-2 | 100 | 2II_E.2 |
| 36 | VIDRL-DN0026 | PX683064 | 2024-02 | 2023-11 | POS | NEG | NEG | 4 | 32 | 27 | DENV-4 | DENV-4 | 84.21 | 4II_A.1 |
| 37 | VIDRL-DN0027 | PX683065 | 2024-02 | 2023-11 | POS | NEG | NEG | 2 | 44 | 38 | DENV-2 | DENV-2 | 74.04 | 2II_E.2 |
| 38 | VIDRL-DN0028 | PX683066 | 2024-02 | 2023-11 | POS | POS | POS | 2 | 39 | 36 | DENV-2 | DENV-2 | 86.85 | 2II_E.2 |
| 39 | VIDRL-DN0029 | PX683067 | 2024-02 | 2023-12 | POS | POS | POS | NA | 30 | 26 | DENV-4 | DENV-4 | 80.16 | 4II_A.1 |
| 40 | VIDRL-DN0030 | PX683068 | 2024-02 | 2024-01 | unknown | unknown | unknown | NA | 40 | 33 | DENV-2 | DENV-2 | 96.92 | 2II_E.2 |
| 41 | VIDRL-DN0031 | PX683069 | 2024-08 | 2024-07 | POS | NEG | POS | 1 | 25 | 19 | DENV-1 | DENV-1 | 98.07 | 1IV_A |
| 42 | VIDRL-DN0032 | PX683070 | 2024-08 | unknown | unknown | unknown | unknown | 2 | 27 | 21 | DENV-2 | DENV-2 | 100 | 2II_E.2 |
| 43 | VIDRL-DN0052 | PX683081 | 2024-08 | unknown | POS | POS | POS | 1 | 38 | 33 | DENV-1 | DENV-1 | 61.83 | 1IV_A |
| 44 | VIDRL-DN0033 | PX683071 | 2024-08 | 2024-06 | POS | NEG | POS | 4 | 35 | 32 | DENV-4 | DENV-4 | 79.96 | 4II_A.1 |
| 45 | VIDRL-DN0034 | PX683072 | 2024-08 | 2024-06 | POS | NEG | NEG | 4 | 27 | 24 | DENV-4 | DENV-4 | 99.97 | 4II_A.1 |
| 46 | VIDRL-DN0053 | PX683082 | 2024-08 | 2024-07 | POS | NEG | NEG | 1 | 22 | 17 | DENV-1 | DENV-1 | 100 | 1IV_A |
| 47 | VIDRL-DN0035 | PX683073 | 2024-08 | 2024-07 | POS | NEG | POS | 1 | 27 | 22 | DENV-1 | DENV-1 | 97.69 | 1IV_A |
| 48 | NA | NA | 2024-08 | 2024-06 | NEG | NEG | POS | 4 | 31 | 28 | DENV-4 | DENV-4 | 59.45 | NA |
| 49 | NA | NA | 2024-08 | unknown | POS | POS | POS | 4 | ND | ND | NA | NA | NA | NA |
| 50 | VIDRL-DN0036 | PX683074 | 2024-08 | unknown | POS | POS | NEG | 2 | 35 | 28 | DENV-2 | DENV-2 | 92.45 | 2II_E.2 |
| 51 | NA | NA | 2025-07 | 2025-03 | POS | NEG | POS | 1 | 44 | 35 | DENV-1 | NA | 43.27 | NA |
| 52 | VIDRL-DN0045 | PX683075 | 2025-07 | 2025-03 | POS | NEG | NEG | 1 | 34 | 28 | DENV-1 | DENV-1 | 91.49 | 1IV_A |
| 53 | NA | NA | 2025-07 | 2025-03 | POS | NEG | NEG | 1 | ND | ND | NA | NA | NA | NA |
| 54 | VIDRL-DN0046 | PX683076 | 2025-07 | 2025-03 | POS | NEG | NEG | 1 | 36 | 29 | DENV-1 | DENV-1 | 85.8 | 1IV_A |
| 55 | VIDRL-DN0047 | PX683077 | 2025-07 | 2025-03 | POS | NEG | NEG | 1 | 36 | 29 | DENV-1 | DENV-1 | 85.69 | 1IV_A |
| 56 | NA | NA | 2025-07 | 2025-03 | unknown | unknown | unknown | 1 | ND | 35 | DENV-1 | NA | 41.75 | NA |
| 57 | VIDRL-DN0048 | PX683078 | 2025-07 | 2024-08 | unknown | unknown | unknown | 4 | 36 | 33 | DENV-4 | DENV-4 | 82.42 | 4II_A.1 |
| 58 | NA | NA | 2025-07 | 2024-08 | unknown | unknown | unknown | 1 | ND | ND | NA | NA | NA | NA |
| 59 | VIDRL-DN0049 | PX683079 | 2025-07 | 2024-08 | unknown | unknown | unknown | 4 | 33 | 30 | DENV-4 | DENV-4 | 85.68 | 4II_A.1 |
| 60 | VIDRL-DN0050 | PX683084 | 2025-07 | 2024-08 | unknown | unknown | unknown | 4 | 40 | 32 | DENV-4 | DENV-4 | 61.79 | 4II_A.1 |
| 61 | NA | NA | 2025-07 | 2024-08 | unknown | unknown | unknown | 4 | ND | ND | NA | NA | NA | NA |
| 62 | VIDRL-DN0051 | PX683080 | 2025-07 | 2024-08 | unknown | unknown | unknown | 2 | 32 | 31 | DENV-2 | DENV-2 | 89.44 | 2II_E.2 |
| 63 | NA | NA | 2025-07 | 2024-08 | unknown | unknown | unknown | 1 | ND | ND | NA | NA | NA | NA |

## References

[1] Samir Bhatt et al. ‘The global distribution and burden of dengue’. In: Nature 496.7446 (2013), pp. 504–507. DOI: 10.1038/nature12060.

[2] Na Tian et al. ‘Dengue Incidence Trends and Its Burden in Major Endemic Regions from 1990 to 2019’. In: Tropical Medicine and Infectious Disease 7.8 (2022), p. 180. ISSN: 2414-6366. URL: https://www.mdpi.com/2414-6366/7/8/180.

[3] E. Togami et al. ‘Epidemiology of dengue reported in the World Health Organization’s West-ern Pacific Region, 2013–2019’. In: Western Pacific Surveillance and Response Journal 14.1 (2023), pp. 1–16. DOI: 10.5365/wpsar.2023.14.1.973.

[4] Rosie J. Matthews et al. ‘Arboviral Disease Outbreaks in the Pacific Islands Countries and Areas, 2014 to 2020: A Systematic Literature and Document Review’. In: Pathogens 11.1 (2022). ISSN: 2076-0817. DOI: 10.3390/pathogens11010074.

[5] National Center for Emerging (NCEZID) and Zoonotic Infectious Diseases. Areas with Risk of Dengue. Web Page. 2025. URL: https://www.cdc.gov/dengue/areas-with-risk/index.html.

[6] Adam T. Craig et al. ‘Enhanced surveillance during a public health emergency in a resource-limited setting: Experience from a large dengue outbreak in Solomon Islands, 2016–17’. In: PLOS ONE 13.6 (2018), e0198487. DOI: 10.1371/journal.pone.0198487.

[7] Kanaporn Poltep et al. ‘Genetic Diversity of Dengue Virus in Clinical Specimens from Bangkok, Thailand, during 2018–2020: Co-Circulation of All Four Serotypes with Multiple Genotypes and/or Clades’. In: Tropical Medicine and Infectious Disease 6.3 (2021), p. 162. ISSN: 2414-6366. URL: https://www.mdpi.com/2414-6366/6/3/162.

[8] Francisco Nogareda et al. ‘Ongoing outbreak of dengue serotype-3 in Solomon Islands, January to May 2013’. In: Western Pacific Surveillance and Response 4.3 (2013). DOI: 10.5365/wpsar.2013.4.2.013.

[9] Adam T. Craig et al. ‘Surveillance strategies for the detection of disease outbreaks in the Pacific islands: meta-analysis of published literature, 2010–2019’. In: Tropical Medicine and International Health 25.8 (2020), pp. 906–918. ISSN: 1360-2276. DOI: 10.1111/tmi.13448.

[10] Wei-Xian Zhang et al. ‘Assessing the global dengue burden: Incidence, mortality, and disability trends over three decades’. In: PLOS Neglected Tropical Diseases 19.3 (2025), e0012932. DOI: 10.1371/journal.pntd.0012932.

[11] Andrew Waleluma Darcy et al. ‘Multiple arboviral infections during a DENV-2 outbreak in Solomon Islands’. In: Tropical Medicine and Health 48.1 (2020), p. 33. ISSN: 1349-4147. DOI: 10.1186/s41182-020-00217-8.

[12] Dong-sheng Li et al. ‘Rapid Displacement of Dengue Virus Type 1 by Type 4, Pacific Region, 2007–2009’. In: Emerging Infectious Disease journal 16.1 (2010), p. 123. ISSN: 1080-6059. DOI: 10.3201/eid1601.091275.

[13] Van-Mai Cao-Lormeau et al. ‘Dengue Virus Type 3, South Pacific Islands, 2013’. In: Emerging Infectious Disease journal 20.6 (2014), p. 1034. ISSN: 1080-6059. DOI: 10.3201/eid2006.131413.

[14] Catherine Inizan et al. ‘Molecular Characterization of Dengue Type 2 Outbreak in Pacific Islands Countries and Territories, 2017–2020’. In: Viruses 12.10 (2020). ISSN: 1999-4915. DOI: 10.3390/v12101081.

[15] Elodie Descloux et al. ‘Dengue 1 Diversity and Microevolution, French Polynesia 2001–2006: Connection with Epidemiology and Clinics’. In: PLOS Neglected Tropical Diseases 3.8 (2009), pp. 1–15. DOI: 0.1371/journal.pntd.0000493.

[16] Marinjho Jonduo et al. ‘Genomic Sequencing of Dengue Virus Strains Associated with Papua New Guinean Outbreaks in 2016 Reveals Endemic Circulation of DENV-1 and DENV-2’. In: The American Journal of Tropical Medicine and Hygiene 107.6 (2022), pp. 1234–1238. DOI: 10.4269/ajtmh.21-1292.

[17] Emma Taylor-Salmon et al. ‘Travel surveillance uncovers dengue virus dynamics and introductions in the Caribbean’. In: medRxiv (2023), p. 2023.11.11.23298412. DOI: 10.1101/2023.11.11.23298412.

[18] Etienne Frumence et al. ‘Dynamics of emergence and genetic diversity of dengue virus in Reunion Island from 2012 to 2022’. In: PLOS Neglected Tropical Diseases 18.5 (2024), e0012184. DOI: 10.1371/journal.pntd.0012184.

[19] The Lancet Regional Health-Western Pacific. ‘Implementing pathogen genomics in the Western Pacific region: evidence is needed’. In: Lancet Reg. Health West. Pac. 47.101134 (2024), p. 101134. DOI: 10.1016/j.lanwpc.2024.101134.

[20] Alyssa T. Pyke et al. ‘First Complete Genome Sequences of Dengue Virus Serotype 2 Strains from the Solomon Islands and Vanuatu’. In: Microbiology Resource Announcements 8.2 (2019), 10.1128/mra.01360–18. DOI: 10.1128/mra.01360-18.

[21] Verity Hill et al. ‘A new lineage nomenclature to aid genomic surveillance of dengue virus’. In: medRxiv (2024), p. 2024.05.16.24307504. DOI: 10.1101/2024.05.16.24307504.

[22] Chantal B. F. Vogels et al. ‘DengueSeq: a pan-serotype whole genome amplicon sequencing protocol for dengue virus’. In: BMC Genomics 25.1 (2024), p. 433. ISSN: 1471-2164. DOI: 10.1186/s12864-024-10350-x.

[23] Catherine Inizan et al. ‘Viral evolution sustains a dengue outbreak of enhanced severity’. In: Emerg. Microbes Infect. 10.1 (2021), pp. 536–544.

[24] Paul F Horwood et al. ‘The Indo-Papuan conduit: a biosecurity challenge for Northern Australia’. In: Aust. N. Z. J. Public Health 42.5 (2018), pp. 434–436. DOI: 10.1111/1753-6405.12808.

[25] PacNet. Epidemic and emerging disease alerts in the Pacific as of 2 September 2025. Press release. 2025. URL: https://reliefweb.int/map/world/epidemic-and-emerging-disease-alerts-pacific-2-september-2025.

[26] World Health Organization. Dengue Guidelines, for Diagnosis, Treatment, Prevention and Control. Geneva: World Health Organization, 2009. URL: https://www.who.int/publications/i/item/9789241547871.

[27] Ivan Aksamentov et al. ‘Nextclade: clade assignment, mutation calling and quality control for viral genomes’. In: J. Open Source Softw. 6.67 (2021), p. 3773. DOI: 10.21105/joss.03773.

[28] John Huddleston et al. ‘Augur: a bioinformatics toolkit for phylogenetic analyses of human pathogens’. In: J. Open Source Softw. 6.57 (2021), p. 2906. DOI: 10.21105/joss.02906.

[29] Kazutaka Katoh and Daron M Standley. ‘MAFFT multiple sequence alignment software version 7: improvements in performance and usability’. In: Mol. Biol. Evol. 30.4 (2013), pp. 772–780. DOI: 10.1093/molbev/mst010.

[30] Bui Quang Minh et al. ‘IQ-TREE 2: New Models and Efficient Methods for Phylogenetic Inference in the Genomic Era’. In: Mol. Biol. Evol. 37.5 (2020), pp. 1530–1534. DOI: 10.1093/molbev/msaa015.

[31] Jaime Huerta-Cepas, François Serra, and Peer Bork. ‘ETE 3: Reconstruction, analysis, and visualization of phylogenomic data’. In: Mol. Biol. Evol. 33.6 (2016), pp. 1635–1638. DOI: 10.1093/molbev/msw046.

